# Socio-economic and demographic determinants of all-cause, main-cause and sub-cause mortality among 45+ adults: Evidence from Longitudinal Ageing Study in India

**DOI:** 10.1101/2022.05.22.22275425

**Authors:** Saddaf Naaz Akhtar, Nandita Saikia

## Abstract

**Background:** Studies on cause-specific mortality among 45+ adults remain unknown in Indian settings. However, understanding the epidemiology of this public health problem can guide policy development for premature and old-age mortality prevention. Therefore, we intend to examine the socio-economic and demographic determinants of all-cause, main-cause and sub-cause mortality among 45+ adults in India.

**Methods:** We adopted the cross-sectional data from the Longitudinal Ageing Study of India (LASI-wave-I) conducted in 2017-18. We performed descriptive, bivariate and multivariate analysis.

**Results:** Females, young-old, middle-old, oldest-old showed lower odds of all-cause, main-cause and sub-cause mortality than males and middle-aged adults. Central region showed significantly greater odds of all-cause mortality risks than Northern region. Christians have lower odds of all-cause mortality risk than Hindus. With the increase in household income, the odds of NCD-related mortality risks also increase. Central (OR=1.54; p<0.01), Eastern (OR=1.28; p<0.01) and Western regions (OR=1.18; p<0.1) have greater odds of non-NCD-related mortality-risks than Northern regions. Urban residence (OR=01.34; p<0.05) has significantly higher odds of CVD-related mortality-risk than rural residence. OBC (OR=0.59; p<0.01) has lower odds of cancer-related mortality risks than general caste. North-eastern region (OR=2.00; p<0.01) has significantly greater odds of diabetes-related mortality risks.

**Conclusions:** The premature and old-age mortality components would help formulate and execute integrated interventions aimed at specific age groups and causes-specific mortality. Medical care, pollution management, environmental control, more involvement in physical activity and a healthy lifestyle could assist in lowering the CVD, cancer & diabetes-related mortality. A new strategy is needed to avoid future deaths and burdens from ageing-related CVD.

**Highlights:**

- This is the first-ever study that provides the socio-economic and demographic factors association of all-cause, main-cause and sub-cause mortality by characteristics among 45+ adults in India.
- Females, young-old, middle-old, and oldest-old showed lower odds of all-cause, main-cause and sub-cause mortality than males and middle-aged adults.
- With the increase in household income, the odds of NCD-related mortality risks also increase.
- Urban residents have significantly higher odds of CVD-related mortality risk than rural residents.
- OBC has lower odds of cancer-related mortality risks than general caste.

## Introduction

According to World Mortality Report (2019), half of the majority of deaths occur among older-adults, even in low-and-middle-income countries (LMICs) (United Nations et al., 2019). Cause-specific mortality among middle-aged and older adults are neglected topic in global health settings, especially in low-and-middle-income countries (LMICs). The World Health Organization report documented that non-communicable (NCDs) related deaths are adversely affected in LMIC, accounting for about 31.4 million deaths, which is three-fourths of global NCDs related deaths (World Health Organization, 2021). It has been observed that even after extended years of infirmity, NCDs frequently result in slow and agonizing deaths. Middle-aged and older adults are more vulnerable to acute health emergencies as a result of the global chronic illness crisis and public health failure to control the growth in highly preventable risk factors.

The term “All-cause mortality” is defined as “the mortality due to any cause of death for a population in a specific time period”. In general, this term is often used by epidemiologists, health professionals and researchers or diseases tracking scientists, to refer to death from any cause. However, anything that causes death is considered to be a cause of death. Hence, all-cause mortality is any cause of death (Basaraba, 2020). Recent study showed that older persons with successful ageing had a 50% lower chance of all-cause mortality (Mao et al., 2022). Older people with diabetes or cardiovascular diseases has strong association with increased risks of all-cause mortality (Chou et al., 2021).

Likewise, noncommunicable diseases (NCDs) are a potential threat to world health and development. Since the early 1980s, NCDs have been on the World Health Organization’s (WHO) agenda, and they have been increased in worldwide (Naghavi et al., 2017; World Health Organization, 1981). As of 2018, the NCD group includes four main diseases by the WHO: cardiovascular diseases (CVD), chronic respiratory diseases (CRD), diabetes, and malignancies, with mental health being added later. There is an urgent need to reduce the burden of NCDs, significant efforts are also required, beginning with the provision of credible epidemiological estimates of NCDs and their drivers in order to properly inform preventative and control initiatives.

However, the most common NCD is cardiovascular disease, which kills 17.9 million people each year, followed by cancer (9.3 million), respiratory disorders (4.1 million), and diabetes (4.1 million) (1.5 million). Over 80% of all premature NCD fatalities are caused by these four disease types (World Health Organization, 2022). Although CVD is preventable and treatable but for the past 20 years, cardiovascular disease (CVD) has remained the top mortality cause in worldwide (World Health Organization, 2020). Since 2000, the number of people dying of CVD has risen by over 2 million, to nearly 9 million in 2019 and it accounts for 16% of all global deaths. Diabetes-related mortality climbed by 70% worldwide between 2000 and 2019, with male deaths increasing by 80% (World Health Organization, 2020). A recent Global burden of diseases report revealed that the global burden of cancer is significant and expanding, with the burden varying according to sociodemographic index which could help to inform global efforts to achieve equitable cancer control (Global Burden of Disease 2019 Cancer Collaboration et al., 2022).

There is a dearth of research on the cause of mortality among adults in any developing country. Therefore, studying the cause of mortality is essential for health professionals and policymakers to understand the different stages of socioeconomic growth for a country. However, various health policies and initiatives programs aim to prevent various diseases and injuries caused among adults (considering high-risk populations) should establish up-to-date and comprehensive information concerning the scope and nature of health issues, their drivers, and how they are affected by such diseases. The socioeconomic drivers in the cause of mortality are required to comprehend the distribution of health problems in any country or society. Though the improvement in the population health can be assessed using a high-quality mortality data source that consists of robust and reliable mortality information by various causes of diseases. In order to enhance the population’s health, there is a need to prioritize health or treatment strategies.

Therefore, we intend to quantify the impact of all-cause, main-cause and sub-cause mortality on socio-economic and demographic determinants among 45+ adults in India.

### Causes-specific mortality at global settings

NCDs are the leading cause of mortality in Europe and it has been found that about 77% of disease burden and 86% of premature mortality are caused due to CVD, diabetes, cancer, and respiratory disease (European commission, 2022). The countries of the United States have documented extremely higher mortality rates due to diabetes and they were located along the Mississippi River’s southern half, as well as in areas of South and North Dakota (Mokdad et al., 2022). Higher mortality occurred due to CVD and cancer among women than men in Sweden from the overall deaths (Rosvall et al., 2006). Another study documented that reported a large contribution of cancer deaths to the excess mortality among low educated middle aged men in France and other Latin countries contrary to Northern Europe countries (Menvielle et al., 2010). Males face more regional mortality differences than females. in the Republic of Ireland (Pringle, 1986). The mortality burden in the Australian population attributable to socioeconomic inequality is large (Turrell & Mathers, 2001). Earlier study in Luxembourg (Mcisaac & Wilkinson, 1997) documented that the lower all-cause mortality rates were associated with a more equitable distribution of income among all ages and both-sexes. A strong association has been observed between the socioeconomic status and mortality risk of a person in a developed society or western society. There has been a plenty of earlier researches evidencing that person from lower socioeconomic backgrounds die at relatively early age or younger age compared than those from better socioeconomic backgrounds (Kunst et al., 1990; Michelozzi et al., 1999; Siskind et al., 2010; Smith et al., 1990; Westerling et al., 1996). A study in Netherland showed that the cancer related mortality are found to be lower among married people (Joung, 1996). Individual social class and area deprivation are both associated with mortality in men in England (Dundas et al., 2014)

Over the last two decades, the burden of NCDs has also increased in Sub-Saharan Africa, owing to rising rates of cardiovascular risk factors such as poor diets, lack of physical activity, hypertension, obesity, diabetes, dyslipidaemia, and air pollution (Bigna & Noubiap, 2019). A study (Wallace & Kulu, 2015) documented that lower death is coexisted with greater CVD-related death, which is the primary cause of death among South Asians (except Bangladeshi women). In China, the CVD-related death is accounting for 36.95% of the total death occurred during the 1974-2015 and greater CVD-related is observed among women but not men(Zhang et al., 2020). A study by (Omar et al., 2019) showed that CVD and diabetes related death are higher among women than men in Malaysia. Females are more influenced by their surroundings, whereas males are more influenced by their socioeconomic status, despite the fact that both have lower mortality rates in rural areas than in metropolitan ones (Allan et al., 2019). A study from Brazil showed that there has been a reduction in the mortality rates due to NCDs from 1990-2017 and the higher premature death due to NCDs are seen among women (Malta et al., 2020). High proportions of “ill-defined” causes of death is observed in seven pacific island countries (Carter et al., 2012). Another study from Kazakhstan showed that the decline in all-cause mortality in less among older adults than the younger adults while the decline is greater among women than men (Davletov et al., 2015).

## Methods

### Data source

Our study has adopted cross-sectional data form the latest Longitudinal Ageing Study in India (LASI, WAVE-I), which was conducted in 2017-18. The LASI is India’s first and the largest amongst the global Health and Retirement Study (HRS) family (IIPS, 2022). It emphasizes on India’s older population’s health, economic, and social well-being. LASI also takes into account the elements that are specific to India, such as its institutional and cultural qualities. The LASI is a large-scale nationwide study of the health, economic, and social factors and repercussions of India’s population aging. The LASI is a national survey of 73,000 older adults aged 45 and up from all Indian states and union territories. For the next 25 years, LASI will be carried out every three years. It is well positioned to assess the impact of policy changes on India’s behavioural outcomes. It is a national milestone in scientific research, allowing for a better understanding of India’s adult health concerns and population aging processes, as well as the development of appropriate evidence-based policies for adults and the elderly. This study gives enough statistical information to evaluate hypotheses in specific subpopulations. LASI data can help scientists enhance their understanding and inform policymakers in India and around the world. For cross-national comparative research studies on aging, the internationally standardized data are useful.

The present study has taken six different effective sample sizes for each cause-specific mortality among older adults aged 45 years and above. The sample size for all-cause is 1,12,590 cases, NCD-related mortality is 1,11,029 cases, non-NCD-related mortality is 1,11,440 cases, CVD-related mortality is 1,10,182 cases, Diabetes-related mortality is 1,10,127 cases, and Cancer-related mortality is 1,10,064 cases respectively. However, we have excluded 4 transgender cases from the sex variable. The study has followed the definition of ICD-10 to define the cause-specific mortality.

### Outcome variables

Our study has included six different dependent variables. All-cause mortality, main-cause mortality includes NCD-related mortality and non-NCD-related mortality, and sub-cause mortality includes CVD-related mortality, Diabetes-related mortality, and Cancer-related mortality.

In the questionnaire, it has been asked that Has any household member died in the last two years (yes or no). If yes, then what was the main cause of death? The list of cause of death information can be found in the Supplementary file. The first dependent variable which we have taken in the present study is all-cause mortality.

1. All-cause mortality (Mortality due to any cause of death): (Yes=1; died due to any cause of death) (No=0; Alive) Since, main-cause mortality comprises four categories are-i) Communicable diseases, ii) Noncommunicable diseases, iii) Injuries and iv) Ill-defined symptoms respectively. In the present study, main-cause included two categories separately are given as:
2. NCD-related mortality: (Yes=1; died due to NCD) (No=0; Alive)
3. Non-NCD-related mortality (includes communicable diseases, injuries and ill-defined symptoms): (Yes=1; died due to non-NCD) (No=0; Alive) Similarly, we have also added sub-cause mortality which has three categories separately are given as:
4. CVD-related mortality: (Yes=1; died due to CVD) (No=0; Alive)
5. Cancer-related mortality: (Yes=1; died due to Cancer) (No=0; Alive)
6. Diabetes related mortality: (Yes=1; died due to Diabetes) (No=0; Alive)

### Independent variables

The present study has several independent variables. Age-groups has four categories such as middle-aged (45-59 years), young-old (60-69 years), middle-old (70-79 years) and oldest-old (80+ years) respectively. Sex has two categories are-male and female. Place includes rural and urban. Regions has six geographical zones-Northern, North-eastern, Central, Eastern, Western and Southern. HH-size includes less than five members and more than or equal to five members. Religion is consisting of four categories such as Hindus, Muslims, Christians and Others (Sikhs, Buddhists, Jains, Parsis etc.). The caste group has four categories, General (no caste), Schedule caste (SC), Schedule Tribe (ST), and Other Backward caste (OBC). Our study has also taken household income variable which comprises of Poorest, Poorer, Middle, Richer and Richest respectively.

### Statistical analysis

Our study has performed descriptive, bivariate, multivariate analysis using R-Studio and STATA (14.0 version) software. The outcomes variables are in the binary (dichotomous) form which are coded as (0= Alive) and (1= Cause-specific mortality); therefore, we have considered binary logistic regression analysis for each outcome variable separately to study the cause-specific mortality-risks of socio-economic and demographic characteristics. In the present study, the binary logistic regression model is used to investigate the effect of predictors on the probability of having mortality among older adults (aged 45+). The expression of binary logistic regression for each outcome variable are as follow:

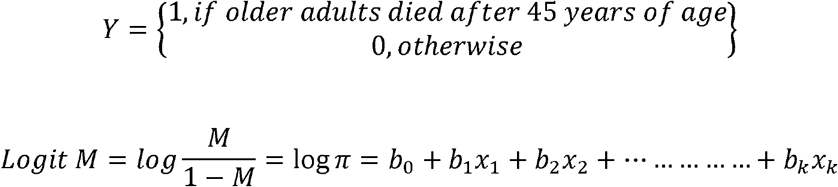

Where, M is the probability that the event Y occurs and coefficient ‘b’ is the factor by which the odds changes with unit increase in independent variable. If ‘b’ is positive, odds ratio (OR) will increase i.e., mortality risk will increase, as this factor will be >1. Contrary to that if ‘b’ <1, then the odds ratio (OR) will decrease i.e., mortality risk will decrease. When ‘b’ is 0, then the factor exponential of ‘b’=1, and therefore, the odds of mortality risk remain unchanged (Ali et al., 2022).

## Results

### Sample distribution of 45+ adults in India

Table 1 presents the sample distributions of the 45+ adults in India by mortality due to all-cause, NCD, Non-NCD, CVD, Cancer and Diabetes with suitable background characteristics using LASI-(Wave-I). Middle-aged adults are showing highest sample percentage followed by young-old and middle-old in all-cause, NCD, Non-NCD, CVD, Cancer and Diabetes while the lowest is observed among the oldest-old. Males have marginally lower sample percentage than females whilst rural residence has higher percentage of 45+ adults in all-cause, NCD, Non-NCD, CVD, Cancer and Diabetes. Interestingly, highest sample percentage of 45+ adults is observed in Southern regions followed by Northern, Eastern and Central whereas North-eastern regions is showing the lowest. More than 5 members have higher percentage of 45+ adults than the less than or equals to five-members. The highest percentage is reflected among Hindus followed by Muslims, Christians and others. Interestingly, OBC is observed as the highest percentage of 45+ adults followed by General and STs while lowest is seen among SCs. However, the poorest is showing the highest percentage of 45+ adults followed by poorer, middle and richer while the lowest is observed among the richest respectively.

**Table 1.** Sample distribution of the 45+ adults in India with suitable background characteristics, 2017-2018.

| Backgrounds | All-Cause of Mortality |  | Main-Cause of Mortality |  |  |  | Sub-Cause of Mortality |  |  |  |  |  |
| --- | --- | --- | --- | --- | --- | --- | --- | --- | --- | --- | --- | --- |
|  | All-Cause |  | NCD |  | Non-NCD |  | CVD |  | Cancer |  | Diabetes |  |
|  | % | N | % | N | % | % | % | N |  |  | % | N |
| **Age groups** |  |  |  |  |  |  |  |  |  |  |  |  |
| Middle aged | 34.04 | 38,324 | 33.72 | 37,440 | 33.83 | 37,696 | 33.57 | 36,986 | 33.53 | 36,906 | 33.56 | 36,954 |
| Young-old | 28.16 | 31,710 | 28.17 | 31,280 | 28.17 | 31,392 | 28.17 | 31,040 | 28.18 | 31,020 | 28.17 | 31,023 |
| Middle-old | 20.76 | 23,373 | 20.88 | 23,179 | 20.84 | 23,222 | 20.94 | 23,070 | 20.95 | 23,055 | 20.94 | 23,062 |
| Oldest-old | 17.04 | 19,183 | 17.23 | 19,130 | 17.17 | 19,130 | 17.32 | 19,086 | 17.34 | 19,083 | 17.33 | 19,088 |
| **Sex** |  |  |  |  |  |  |  |  |  |  |  |  |
| Male | 49.39 | 55,606 | 49.1 | 54,512 | 49.3 | 54,935 | 49 | 53,994 | 49.02 | 53,948 | 49.03 | 53,994 |
| Female | 50.61 | 56,984 | 50.9 | 56,517 | 50.7 | 56,505 | 51 | 56,188 | 50.98 | 56,116 | 50.97 | 56,133 |
| **Place** |  |  |  |  |  |  |  |  |  |  |  |  |
| Rural | 66.16 | 74,491 | 66.13 | 73,418 | 66.22 | 73,796 | 66.15 | 72,890 | 66.18 | 72,839 | 66.18 | 72,883 |
| Urban | 33.84 | 38,099 | 33.87 | 37,611 | 33.78 | 37,644 | 33.85 | 37,292 | 33.82 | 37,225 | 33.82 | 37,244 |
| **Regions** |  |  |  |  |  |  |  |  |  |  |  |  |
| Northern | 19.21 | 21,624 | 19.25 | 21,375 | 19.16 | 21,355 | 19.23 | 21,186 | 19.22 | 21,151 | 19.21 | 21,155 |
| North-Eastern | 13.55 | 15,257 | 13.6 | 15,104 | 13.53 | 15,077 | 13.56 | 14,944 | 13.59 | 14,953 | 13.62 | 14,999 |
| Central | 14.31 | 16,109 | 14.25 | 15,819 | 14.33 | 15,969 | 14.26 | 15,710 | 14.27 | 15,705 | 14.26 | 15,706 |
| Eastern | 18.06 | 20,329 | 18.03 | 20,015 | 18.09 | 20,156 | 18.05 | 19,885 | 18.05 | 19,869 | 18.05 | 19,881 |
| Western | 13.84 | 15,584 | 13.84 | 15,361 | 13.84 | 15,426 | 13.85 | 15,255 | 13.83 | 15,223 | 13.82 | 15,221 |
| Southern | 21.04 | 23,687 | 21.04 | 23,355 | 21.05 | 23,457 | 21.06 | 23,202 | 21.05 | 23,163 | 21.03 | 23,165 |
| **HH-size** |  |  |  |  |  |  |  |  |  |  |  |  |
| <=5 member | 33 | 37,155 | 32.78 | 36,391 | 32.82 | 36,579 | 32.6 | 35,922 | 32.53 | 35,808 | 32.57 | 35,871 |
| >5members | 67 | 75,435 | 67.22 | 74,638 | 67.18 | 74,861 | 67.4 | 74,260 | 67.47 | 74,256 | 67.43 | 74,256 |
| **Religion** |  |  |  |  |  |  |  |  |  |  |  |  |
| Hindus | 71.79 | 80,828 | 71.7 | 79,612 | 71.79 | 80,006 | 71.71 | 79,012 | 71.69 | 78,910 | 71.69 | 78,950 |
| Muslims | 13.73 | 15,460 | 13.75 | 15,271 | 13.74 | 15,317 | 13.77 | 15,173 | 13.77 | 15,156 | 13.77 | 15,160 |
| Christians | 9.75 | 10,975 | 9.8 | 10,877 | 9.74 | 10,857 | 9.78 | 10,776 | 9.8 | 10,787 | 9.81 | 10,802 |
| Others | 4.73 | 5,327 | 4.75 | 5,269 | 4.72 | 5,260 | 4.74 | 5,221 | 4.73 | 5,211 | 4.74 | 5,215 |
| **Caste** |  |  |  |  |  |  |  |  |  |  |  |  |
| General | 27.38 | 30,831 | 27.43 | 30,456 | 27.34 | 30,471 | 27.41 | 30,198 | 27.4 | 30,162 | 27.39 | 30,164 |
| SC | 17.32 | 19,503 | 17.3 | 19,205 | 17.32 | 19,302 | 17.29 | 19,055 | 17.3 | 19,037 | 17.29 | 19,041 |
| ST | 17.91 | 20,165 | 17.94 | 19,922 | 17.93 | 19,981 | 17.94 | 19,765 | 17.97 | 19,775 | 17.98 | 19,804 |
| OBC | 37.38 | 42,091 | 37.33 | 41,446 | 37.41 | 41,686 | 37.36 | 41,164 | 37.33 | 41,090 | 37.34 | 41,118 |
| **HH-Income** |  |  |  |  |  |  |  |  |  |  |  |  |
| Poorest | 24.11 | 27,140 | 24.15 | 26,810 | 24.2 | 26,965 | 24.21 | 26,670 | 24.22 | 26,660 | 24.23 | 26,687 |
| Poorer | 21.69 | 24,425 | 21.75 | 24,151 | 21.72 | 24,205 | 21.76 | 23,977 | 21.78 | 23,973 | 21.78 | 23,986 |
| Middle | 19.82 | 22,311 | 19.8 | 21,983 | 19.82 | 22,087 | 19.8 | 21,821 | 19.8 | 21,793 | 19.8 | 21,808 |
| Richer | 18.14 | 20,422 | 18.11 | 20,111 | 18.11 | 20,178 | 18.1 | 19,943 | 18.08 | 19,905 | 18.07 | 19,904 |
| Richest | 16.25 | 18,292 | 16.19 | 17,974 | 16.16 | 18,005 | 16.13 | 17,771 | 16.11 | 17,733 | 16.11 | 17,742 |
| **Total** | **100** | **1,12,590** | **100** | **1,11,029** | **100** | **1,11,440** | **100** | **1,10,182** | **100** | **1,10,064** | **100** | **1,10,127** |

**Source:** Authors' own calculation using LASI (Wave-1)
**Notes:** We have included communicable diseases, Injuries and ill-defined cause of death as a non-NCD related mortality category.
**Abbreviations:** CVD-Cardiovascular diseases; HH- household; SC-Schedule Caste; ST-Schedule Tribe; OBC-Other Backward Caste

### NCD-related mortality among 45+ adults in India

Figure 1 presents the percentage share of NCD-related mortality among 45+ adults in India with suitable socio-economic and demographic characteristics. The percentage of NCD-related mortality is found to be 37.57% among 45+ adults in India. The middle-aged adults (39.11%) are showing the highest percentage of NCD-related mortality followed by middle-old (37.1%) and young-old (36.37%) and lowest is observed among oldest-old adults (32.06%). Females (42%) are showing higher percentage share of NCD-related mortality than males. Urban residence (40.06%) is found to have greater percentage share of NCD-related mortality than rural residence. The Northern regions (52.48%) has the highest percentage share of NCD-related mortality while lowest is seen in Southern regions (30.73%). Household size with more than five-members are showing marginally equal percentage share of NCD-related mortality compared to household size (<=5 members). Other religion group (48.88%) is showing the highest percentage share of NCD-related mortality followed by Christians (45.06%) and Muslims (40.04%) and the lowest is seen among Hindus (36.62%) respectively. However, the highest percentage share of NCD-related mortality is found to be among General caste group (40.19%) and lowest is seen among STs (34.64%). The percentage share of NCD-related mortality is found to be greatest among the poorer (43.08%) followed by Richest (39.54%), Middle (38.36%) and Richer (36.19%) and the lowest is observed among poorest (31.64%) respectively.

**Figure 1.**
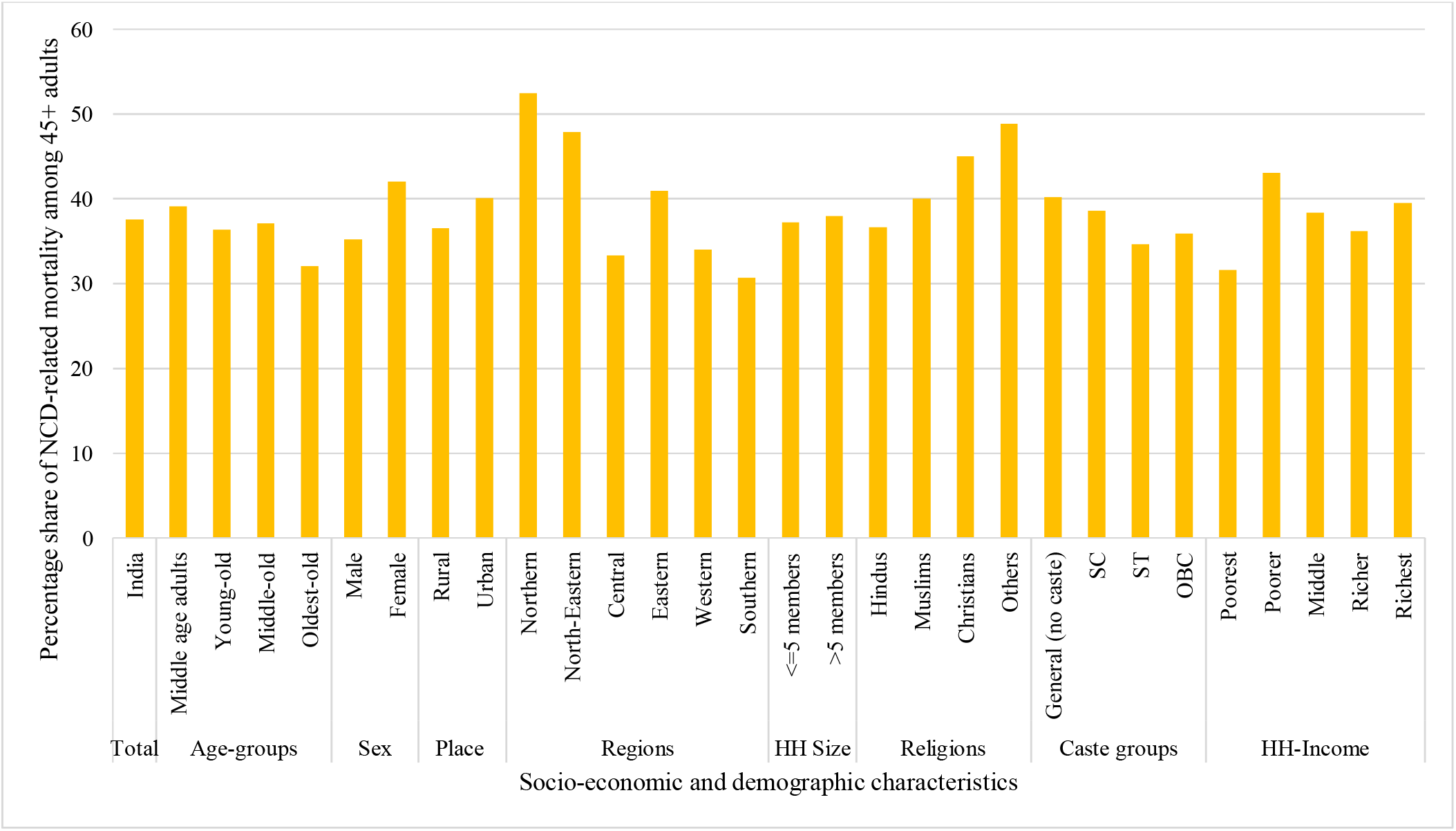
Percent distribution of main-cause (NCD-related death) of mortality among 45+ adults in India. **Source:** Authors’ own calculation using LASI (Wave-1) **Notes:** We have included communicable diseases, Injuries and ill-defined cause of death as a non-NCD related mortality category. **Abbreviations:** HH-household; SC-Schedule Caste; ST-Schedule Tribe; OBC-Other Backward Caste

### Sub-cause (CVD, Cancer and Diabetes) mortality among 45+ adults in India

Figure 2 presents the percentage share of sub-cause (CVD, Cancer and Diabetes) of mortality among 45+ adults in India with suitable socio-economic and demographic characteristics. The overall percentage share of CVD-related mortality among 45+ adults is 10.5% in India, while cancer-related mortality is found to be 6.41% and diabetes-related mortality has 5.65%. Middle-old and females have the highest percentage share of CVD-related and cancer-related mortality but middle-aged adults and males have highest percentage share of diabetes-related mortality. Nevertheless, urban residence is showing higher percentage share of CVD-related mortality while rural residence is found to have greater cancer & diabetes-related mortality. Interestingly, Northern region has the highest percentage share of CVD-related mortality and North-eastern is showing greater cancer & diabetes-related mortality. Household size (<=5 members) has greater percentage share of CVD-related mortality while household size with (>5 members) has higher percentage share of cancer & diabetes-related mortality. Hindus are showing highest percentage share of CVD-related mortality but Christians are found to have greater percentage share of cancer & diabetes-related mortality. General caste is showing greater percentage share of CVD-related mortality whereas STs has higher cancer-related mortality but SCs has greater diabetes-related mortality respectively. However, Richest are showing higher percentage share of CVD-related mortality.

**Figure 2.**
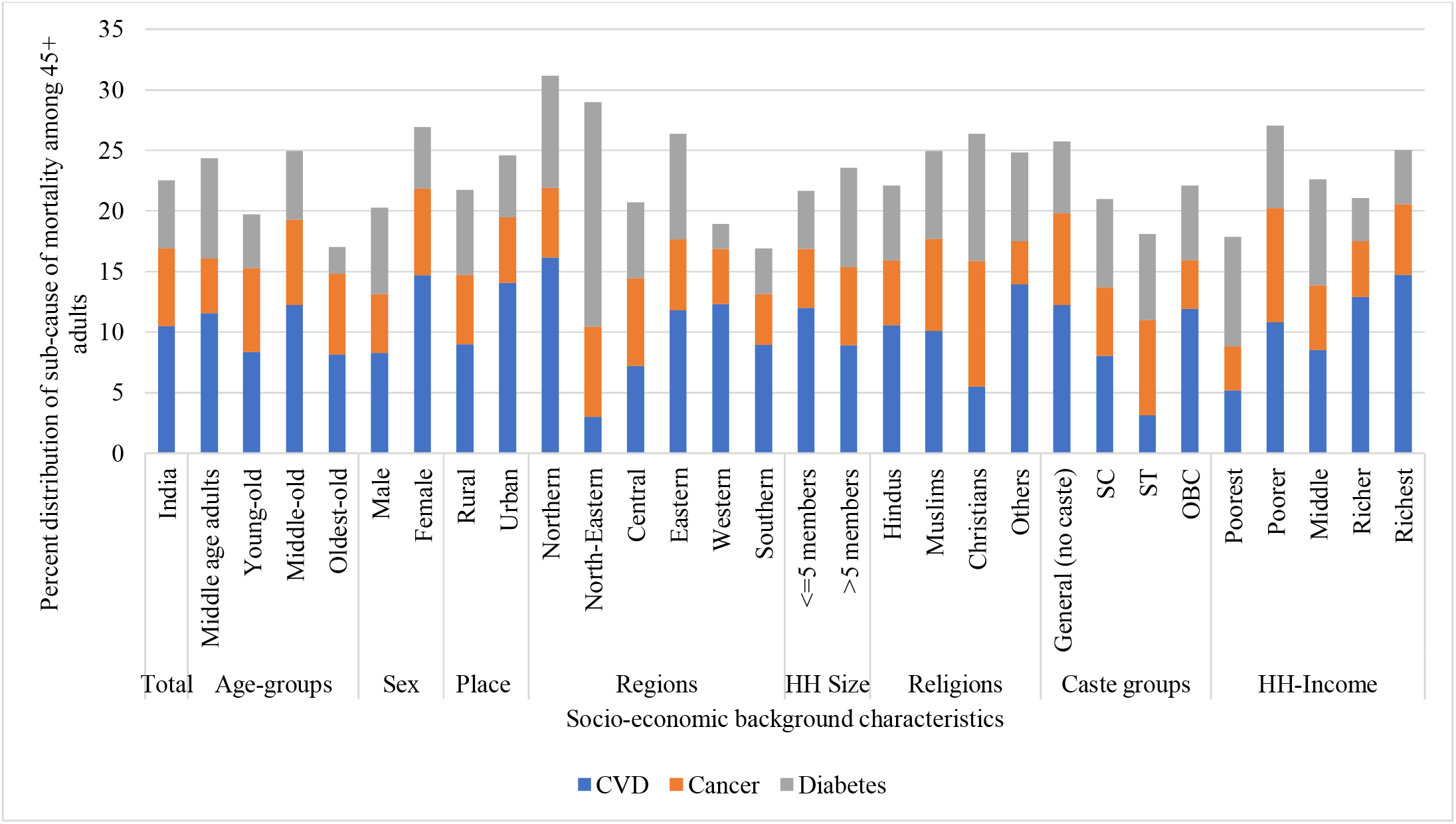
Percentage share of sub-cause (CVD, Cancer and Diabetes) of mortality among 45+ adults in India. **Source:** Authors’ own calculation using LASI (Wave-1) **Notes:** We have only included top three sub-cause of mortality, i.e., CVD, Cancer and Diabetes. **Abbreviations:** CVD-Cardiovascular diseases; HH-household; SC-Schedule Caste; ST-Schedule Tribe; OBC-Other Backward Caste

### Risk factors of mortality due to all-cause, NCD and non-NCD among 45+adults in India

Table 2 shows the regression results of mortality due to all-cause (Model-1), NCD (Model-2), and Non-NCD (Model-3) among 45+ adults with suitable background characteristics.

**Table 2.**
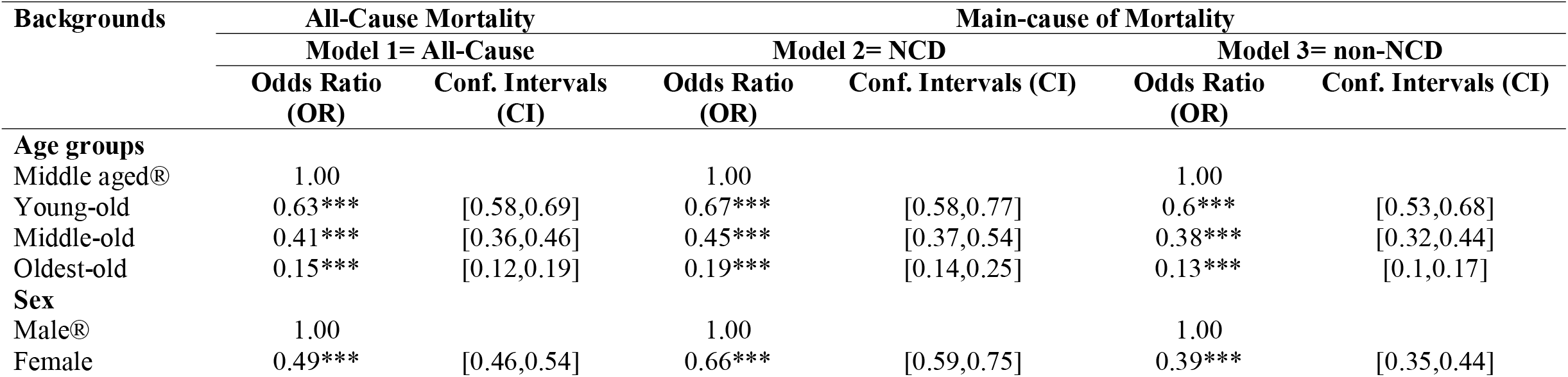
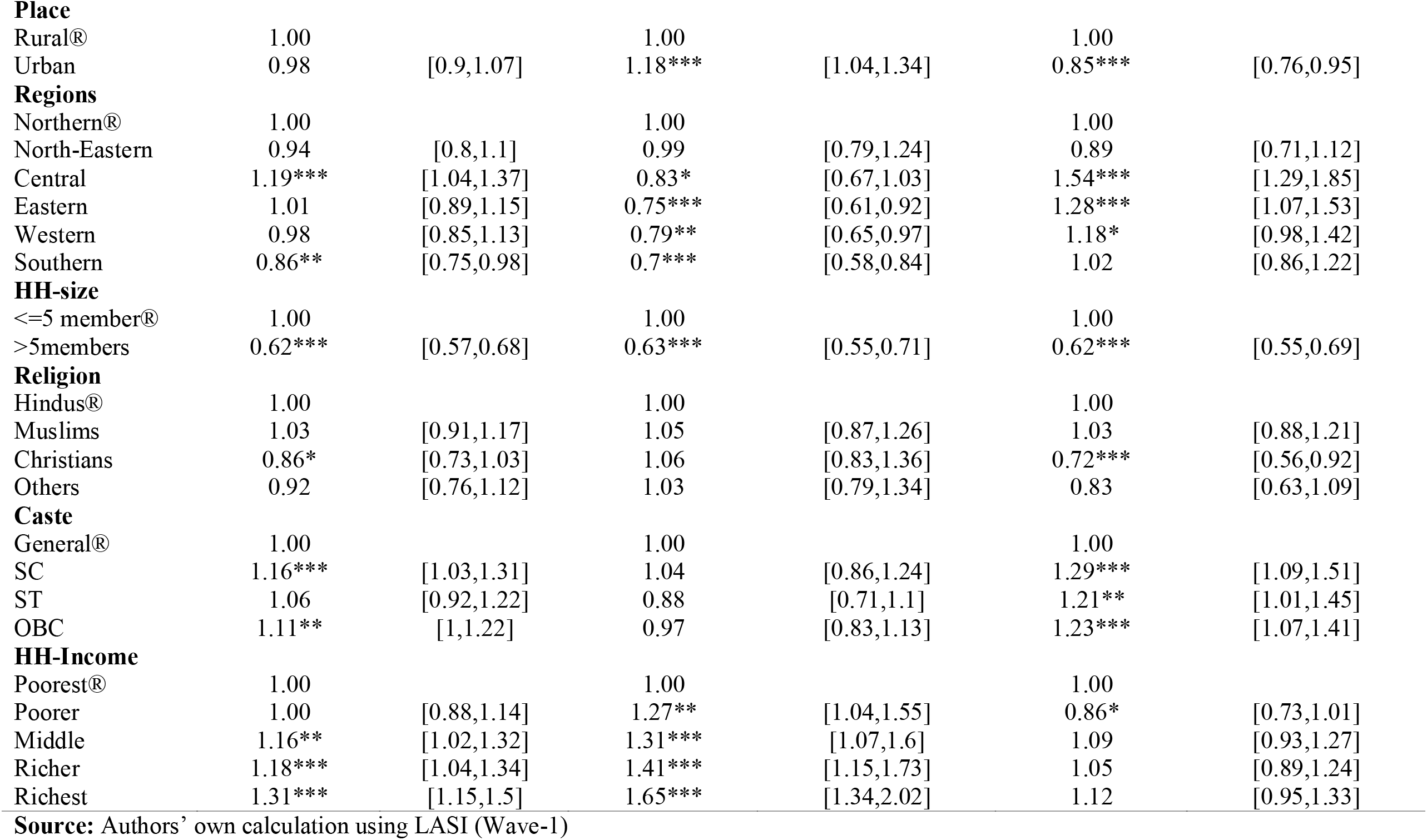
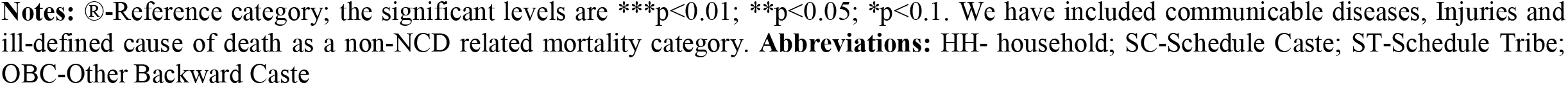
Binary logistics regression results of all-cause and main-cause of mortality among 45+ adults in India.

| Backgrounds | All-Cause Mortality |  | Main-cause of Mortality |  |  |  |
| --- | --- | --- | --- | --- | --- | --- |
|  | Model 1= All-Cause |  | Model 2= NCD |  | Model 3= non-NCD |  |
|  | Odds Ratio (OR) | Conf. Intervals (CI) | Odds Ratio (OR) | Conf. Intervals (CI) | Odds Ratio (OR) | Conf. Intervals (CI) |
| **Age groups** |  |  |  |  |  |  |
| Middle aged® | 1.00 |  | 1.00 |  | 1.00 |  |
| Young-old | 0.63*** | [0.58,0.69] | 0.67*** | [0.58,0.77] | 0.6*** | [0.53,0.68] |
| Middle-old | 0.41*** | [0.36,0.46] | 0.45*** | [0.37,0.54] | 0.38*** | [0.32,0.44] |
| Oldest-old | 0.15*** | [0.12,0.19] | 0.19*** | [0.14,0.25] | 0.13*** | [0.1,0.17] |
| **Sex** |  |  |  |  |  |  |
| Male® | 1.00 |  | 1.00 |  | 1.00 |  |
| Female | 0.49*** | [0.46,0.54] | 0.66*** | [0.59,0.75] | 0.39*** | [0.35,0.44] |
| **Place** |  |  |  |  |  |  |
| Rural® | 1.00 |  | 1.00 |  | 1.00 |  |
| Urban | 0.98 | [0.9,1.07] | 1.18*** | [1.04,1.34] | 0.85*** | [0.76,0.95] |
| **Regions** |  |  |  |  |  |  |
| Northern® | 1.00 |  | 1.00 |  | 1.00 |  |
| North-Eastern | 0.94 | [0.8,1.1] | 0.99 | [0.79,1.24] | 0.89 | [0.71,1.12] |
| Central | 1.19*** | [1.04,1.37] | 0.83* | [0.67,1.03] | 1.54*** | [1.29,1.85] |
| Eastern | 1.01 | [0.89,1.15] | 0.75*** | [0.61,0.92] | 1.28*** | [1.07,1.53] |
| Western | 0.98 | [0.85,1.13] | 0.79** | [0.65,0.97] | 1.18* | [0.98,1.42] |
| Southern | 0.86** | [0.75,0.98] | 0.7*** | [0.58,0.84] | 1.02 | [0.86,1.22] |
| **HH-size** |  |  |  |  |  |  |
| <=5 member® | 1.00 |  | 1.00 |  | 1.00 |  |
| >5members | 0.62*** | [0.57,0.68] | 0.63*** | [0.55,0.71] | 0.62*** | [0.55,0.69] |
| **Religion** |  |  |  |  |  |  |
| Hindus® | 1.00 |  | 1.00 |  | 1.00 |  |
| Muslims | 1.03 | [0.91,1.17] | 1.05 | [0.87,1.26] | 1.03 | [0.88,1.21] |
| Christians | 0.86* | [0.73,1.03] | 1.06 | [0.83,1.36] | 0.72*** | [0.56,0.92] |
| Others | 0.92 | [0.76,1.12] | 1.03 | [0.79,1.34] | 0.83 | [0.63,1.09] |
| **Caste** |  |  |  |  |  |  |
| General® | 1.00 |  | 1.00 |  | 1.00 |  |
| SC | 1.16*** | [1.03,1.31] | 1.04 | [0.86,1.24] | 1.29*** | [1.09,1.51] |
| ST | 1.06 | [0.92,1.22] | 0.88 | [0.71,1.1] | 1.21** | [1.01,1.45] |
| OBC | 1.11** | [1,1.22] | 0.97 | [0.83,1.13] | 1.23*** | [1.07,1.41] |
| **HH-Income** |  |  |  |  |  |  |
| Poorest® | 1.00 |  | 1.00 |  | 1.00 |  |
| Poorer | 1.00 | [0.88,1.14] | 1.27** | [1.04,1.55] | 0.86* | [0.73,1.01] |
| Middle | 1.16** | [1.02,1.32] | 1.31*** | [1.07,1.6] | 1.09 | [0.93,1.27] |
| Richer | 1.18*** | [1.04,1.34] | 1.41*** | [1.15,1.73] | 1.05 | [0.89,1.24] |
| Richest | 1.31*** | [1.15,1.5] | 1.65*** | [1.34,2.02] | 1.12 | [0.95,1.33] |

**Source:** Authors' own calculation using LASI (Wave-1)
**Notes:** ®-Reference category; the significant levels are \*\*\*p<0.01; \*\*p<0.05; \*p<0.1. We have included communicable diseases, Injuries and ill-defined cause of death as a non-NCD related mortality category. **Abbreviations:** HH- household; SC-Schedule Caste; ST-Schedule Tribe; OBC-Other Backward Caste

In Model-1, the young-old (OR=0.63; p<0.01), middle-old (OR=0.41; p<0.01) and oldest-old (OR=0.15; p<0.01) are showing significantly lower odds of all-cause mortality risks compared to the middle-aged adults. Females (OR=0.49; p<0.01) have significantly lower odds of all-cause mortality risk than males. The Central region shows significantly greater odds of all-cause mortality risks than the Northern region, while the lower is observed in the Southern region (OR=0.86; p<0.05). Lower odds are seen among household size with more than five members (OR=0.49; p<0.01) than in households with less than or equal to five members. Christians (OR=0.86; p<0.1) show lower odds of all-cause mortality risk than Hindus. However, SC (OR=1.16; p<0.01) and OBC (OR=1.11; p<0.05) show significantly higher odds of all-cause mortality risks than the General caste. Interestingly, greater odds of all-cause mortality risk are reflected among the middle-income group (OR=1.16; p<0.05), richer (OR=1.18; p<0.01) and richer (OR=1.31; p<0.01) compared to poorest.

Likewise, in Model-2, the young-old (OR=0.67; p<0.01), middle-old (OR=0.45; p<0.01), oldest-old (OR=0.19; p<0.01) and females (OR=0.66; p<0.01) are also showing significantly lower odds of NCD-related mortality risks. Urban residence (OR=1.18; p<0.01), is found to be significantly greater odds of NCD-related mortality risks compared to rural. Interestingly, significantly lower odds of NCD-related mortality risks are observed in the Central (OR=0.83; p<0.1), Eastern (OR=0.75; p<0.01), Western (OR=0.79; p<0.05), and Southern regions (OR=0.67; p<0.01) than Northern region. Household size with more than five members (OR=0.63; p<0.01) are showing lower odds of NCD-related mortality risks which is similar to all-cause mortality. With increase in the household income, the odds of NCD-related mortality risks also get increases, i.e., Poorer (OR=1.27; p<0.05), Middle (OR=1.31; p<0.01), Richer (OR=1.41; p<0.01), and Richest (OR=1.65; p<0.01) are showing higher odds of NCD-related mortality risks.

Furthermore, in Model-3, the young-old (OR=0.6; p<0.01), middle-old (OR=0.38; p<0.01), oldest-old (OR=0.13; p<0.01) and females (OR=0.39; p<0.01) are also showing significantly lower odds of non-NCD-related mortality risks which are similar to all-cause and NCDs-related mortality. But significantly lower odds of non-NCD-related mortality risk is observed in urban residence (OR=0.85; p<0.01) than rural. Central (OR=1.54; p<0.01), Eastern (OR=1.28; p<0.01) and Western regions (OR=1.18; p<0.1) are showing greater odds of non-NCD-related mortality risks than Northern regions. Household size with more than five members (OR=0.62; p<0.01) are showing lower odds of non-NCD-related mortality risks which is similar to all-cause & NCDs-related mortality. Christians (OR=0.72; p<0.01) are showing significantly lower odd of non-NCD-related mortality risks than Hindus which is similar to the all-cause mortality. However, SC (OR=1.29; p<0.01), ST (OR=1.21; p<0.05) and OBC (OR=1.23; p<0.01) are showing significantly greater odds of non-NCD-related mortality risks than General caste. Lastly, the lower odds of non-NCD-related mortality risk is seen only among poorer income group than poorest respectively.

### Risk factors of mortality due to CVD, Cancer and Diabetes among 45+adults in India

Table 3 indicates the regression results of mortality due to CVD (Model-1), Cancer (Model-2), and Diabetes (Model-3) among 45+ adults with suitable background characteristics.

**Table 3.** Binary logistics regression results of sub-cause (CVD, Diabetes and Cancer) of mortality among 45+ adults in India.

| Backgrounds | Sub-Cause of Mortality |  |  |  |  |  |
| --- | --- | --- | --- | --- | --- | --- |
|  | Model 1= CVD |  | Model 2= Cancer |  | Model 3= Diabetes |  |
|  | Odds Ratio (OR) | Conf. Intervals (CI) | Odds Ratio (OR) | Conf. Intervals (CI) | Odds Ratio (OR) | Conf. Intervals (CI) |
| **Age groups** |  |  |  |  |  |  |
| Middle aged® | 1.00 |  | 1.00 |  | 1.00 |  |
| Young-old | 0.62*** | [0.47,0.82] | 0.84 | [0.6,1.17] | 0.56*** | [0.41,0.76] |
| Middle-old | 0.46*** | [0.32,0.65] | 0.49*** | [0.31,0.78] | 0.39*** | [0.26,0.58] |
| Oldest-old | 0.13*** | [0.07,0.26] | 0.15*** | [0.06,0.34] | 0.16*** | [0.09,0.30] |
| **Sex** |  |  |  |  |  |  |
| Male® | 1.00 |  | 1.00 |  | 1.00 |  |
| Female | 0.91 | [0.72,1.14] | 0.66*** | [0.49,0.89] | 0.56*** | [0.43,0.73] |
| **Place** |  |  |  |  |  |  |
| Rural® | 1.00 |  | 1.00 |  | 1.00 |  |
| Urban | 1.34** | [1.05,1.7] | 1.09 | [0.79,1.5] | 1.13 | [0.86,1.49] |
| **Regions** |  |  |  |  |  |  |
| Northern® | 1.00 |  | 1.00 |  | 1.00 |  |
| North-Eastern | 0.46*** | [0.26,0.8] | 0.72 | [0.4,1.31] | 2.00*** | [1.31,3.07] |
| Central | 0.76 | [0.5,1.16] | 1.15 | [0.7,1.9] | 0.87 | [0.53,1.40] |
| Eastern | 0.65** | [0.44,0.97] | 0.64* | [0.38,1.07] | 0.78 | [0.49,1.22] |
| Western | 0.94 | [0.66,1.36] | 0.63* | [0.37,1.09] | 0.48*** | [0.28,0.83] |
| Southern | 0.78 | [0.55,1.1] | 0.69 | [0.42,1.12] | 0.60** | [0.38,0.95] |
| **HH-size** |  |  |  |  |  |  |
| <=5 member® | 1.00 |  | 1.00 |  | 1.00 |  |
| >5members | 0.67*** | [0.52,0.86] | 0.65*** | [0.47,0.9] | 0.68*** | [0.51,0.89] |
| **Religion** |  |  |  |  |  |  |
| Hindus® | 1.00 |  | 1.00 |  | 1.00 |  |
| Muslims | 1.2 | [0.86,1.68] | 1.35 | [0.87,2.1] | 1.10 | [0.74,1.65] |
| Christians | 0.82* | [0.45,1.49] | 1.75** | [0.99,3.07] | 0.96 | [0.59,1.58] |
| Others | 1.15 | [0.7,1.88] | 0.92 | [0.45,1.86] | 0.93 | [0.51,1.67] |
| **Caste** |  |  |  |  |  |  |
| General® | 1.00 |  | 1.00 |  | 1.00 |  |
| SC | 1.03 | [0.72,1.46] | 0.93 | [0.59,1.45] | 0.94 | [0.62,1.44] |
| ST | 0.63* | [0.38,1.03] | 0.81 | [0.48,1.37] | 1.10 | [0.70,1.72] |
| OBC | 1.07 | [0.8,1.42] | 0.59*** | [0.4,0.89] | 0.94 | [0.66,1.33] |
| **HH-Income** |  |  |  |  |  |  |
| Poorest® | 1.00 |  | 1.00 |  | 1.00 |  |
| Poorer | 1.3 | [0.82,2.05] | 1.76** | [1.05,2.96] | 1.06 | [0.72,1.58] |
| Middle | 1.74*** | [1.13,2.69] | 1.39 | [0.8,2.41] | 0.95 | [0.63,1.43] |
| Richer | 2.26*** | [1.48,3.45] | 1.76** | [1.03,3.02] | 0.77 | [0.50,1.20] |
| Richest | 2.49*** | [1.62,3.83] | 2.07*** | [1.2,3.55] | 1.13 | [0.74,1.71] |

**Source:** Authors' own calculation using LASI (Wave-1)
**Notes:** ®-Reference category; the significant levels are \*\*\*p<0.01; \*\*p<0.05; \*p<0.1. **Abbreviations:** CVD-cardiovascular diseases; HH-household; SC-Schedule Caste; ST-Schedule Tribe; OBC-Other Backward Caste

In Model-1, the young-old (OR=0.62; p<0.01), middle-old (OR=0.46; p<0.01) and oldest-old (OR=0.13; p<0.01) are showing significantly lower odds of CVD-related mortality risks compared to the middle-aged adults. Urban residence (OR=01.34; p<0.05) has significantly greater odds of CVD-related mortality risk than rural residence. The North-eastern (OR=0.46; p<0.01) and Eastern regions (OR=0.65; p<0.05) are found to have lower odds of CVD-related mortality risks than the Northern region. Household size with more than five members (OR=0.67; p<0.01), Christians (OR=0.82; p<0.1) and ST (OR=0.63; p<0.1) are showing lower odds of CVD-related mortality risks. However, middle-income (OR=1.74; p<0.01), richer (OR=2.26; p<0.01) and richest (OR=2.49; p<0.01) are found to have significantly greater CVD-related mortality risks than the poorest.

While in Model-2, the lower odds of cancer-related mortality risks are observed among middle-old (OR=0.49; p<0.01), oldest-old (OR=0.15; p<0.01) and females (OR=0.66; p<0.01). The Eastern (OR=0.64; p<0.1) and Western regions (OR=0.63; p<0.1) are also showing lower odds of cancer-related mortality risks compared to Northern regions. Lower mortality risk is reflected among the household size with more than five members (OR=0.65; p<0.01). Muslims (OR=1.75; p<0.05) are found to have greater odds of cancer-related mortality risks than Hindus whereas OBC (OR=0.59; p<0.01) is showing lower odds of cancer-related mortality risks compared to General caste. Meanwhile, greater odds of cancer-related mortality risks are seen among poorer (OR=1.76; p<0.05), richer (OR=1.76; p<0.05) and richest (OR=2.07; p<0.01) than the poorest.

However, in Model-3, the lower odds of diabetes-related mortality risks are observed among young-old (OR=0.56; p<0.01), middle-old (OR=0.39; p<0.01), oldest-old (OR=0.16; p<0.01) and females (OR=0.56; p<0.01). The North-eastern region (OR=2.00; p<0.01) is showing the significantly greater odds of diabetes-related mortality risks while Western (OR=0.48; p<0.01) and Southern (OR=0.60; p<0.05) are showing lower odds of diabetes-related mortality risks than the Northern region. Likewise, lower mortality risk is reflected among the household size with more than five members (OR=0.68; p<0.01).

## Discussion

Mortality measures by cause are essential contributors to population health assessment, policy, and research (Rao & Gupta, 2020). For a coherent public health strategy, cause-specific research on socioeconomic patterns in mortality is also urgently required in any developing nations. However, cause-specific mortality are a reflection of numerous impacts and processes that range from medical advances, changes in lifestyles and nutrition to epidemics and aging. In spite of the evident differences between the past experience of different countries, it is still reasonable to expect that some of these processes will be present in all countries due to their universal character, e.g., aging (Arnold & Glushko, 2021). Therefore, our study has investigated the all-cause, main-cause (NCD and non-NCD-related mortality) and sub-cause (CVD, Cancer and Diabetes-related) mortality among older adults in India.

Our finding revealed that males are significantly at a greater risk of all-cause, main-cause and sub-cause mortality risks than females, though historically, similar studies have been well documented in the previous studies (Helweg-Larsen & Juel, 2000; Koskenvuo et al., 1986; Rosella et al., 2016; Tiainen et al., 2013; Waldron, 1993; Wingard, 1982). However, gender disparities in mortality studies are little understood among 45+ adults in Indian context. Given the fast rise in the number of nonagenarians, greater data on mortality predictors in 45+ age groups could aid in better understanding population ageing and promoting health and functioning among the elderly. Interestingly, our study has revealed that middle-aged adults have higher all-cause, NCD & non-NCD-related mortality risks compared to older adults. However, a recent study by Jinyuan Qi (Qi, 2020) well documented that lower premature NCD death and higher decline of premature NCD death is observed in higher-income countries, while higher premature NCD mortality and lower decline of premature death is seen in lower-income countries.

Though, India is currently undergoing a massive surge of demographic change, leading to a substantial increase in the population of elderly people aged 60 and up. The rising prevalence of NCD evidenced with the problem of population ageing. Hence, the demographic and epidemiological transition has shifted a large portion of disease burden to the middle-aged and older populations. Our study revealed that higher-socioeconomic status has greater NCD & CVD & Cancer-related mortality risks among 45+ adults in India while the previous studies in China (Wang & Wang, 2020; Yu et al., 2017) reflected opposite finding from our study. They showed that as the level of economic development rises, more patients with NCDs will be able to afford the cost of treatment and would seek medical treatment. As medical conditions improve, NCDs will be effectively treated, thus the mortality would be reduced. Meanwhile, recent studies have found that rapid urbanization in India is causing a shift in lifestyle among urban residence, with poor dietary habits mainly depending on fast-or junk food and also lower participation in the physical activity leading to higher prevalence of chronic morbidities among older adults (Chauhan, Patel, et al., 2022; Chauhan, Srivastava, et al., 2022; Khan et al., 2022). Hence, our finding revealed that 45+ adults belonging to urban residences have greater odds of NCD & CVD-related mortality risks than rural.

India is one of the world’s most hierarchical societies where health and social disadvantage are inextricably connected (Vyas et al., 2022). However, in today’s India, concerns about the mortality and health of India’s underprivileged social groups are still pressing and recent studies have highlighted that SC & ST have much shorter life expectancies than the upper caste (general caste) and these enormous differences have lasted nearly for two decades, from 1997 to 2016 (Gupta & Sudharsanan, 2022; Vyas et al., 2022). They have also indicated that mortality disparities in India are not just a childhood phenomenon, but are also increasing more and more problems among older populations too (Gupta & Sudharsanan, 2022). Another recent study evident that social disadvantage groups have higher share of hospitalizations due to communicable diseases than NCDs (Akhtar & Saikia, 2022) while other study showed that social disadvantage groups have higher premature mortality in India than general caste (Kumari & Mohanty, 2020).Thus, our finding evidencing ST, SC and OBC have significantly greater odds of non-NCD-related mortality risks than the general caste.

Besides that, our study has also found that Christians have higher odds of cancer-related mortality risk but lower odds of all-cause and non-NCD-related mortality risks among 45+adults compared to Hindus. Though, higher life expectancy at birth is observed among Christians compared to Hindus and Muslims in India (Kumari & Mohanty, 2020) while previous study showed that Christians have higher lungs-cancer-related mortality risk than Hindus (Gajalakshmi et al., 1996). However, earlier research suggested that the effects of religion on health and mortality risk had been difficult to predict but also highlighted that religion has strong impact on cancer-related mortality (Dwyer et al., 1990; Kretzler et al., 2020).

Previous study have suggested that the effect of family size on mortality varies from countries to countries, depending on the economic conditions whether growing up in a large family may be considered as a disadvantage in a country context or not (Baranowska-Rataj et al., 2017). Though, another study (Feng et al., 2017) has depicted higher mortality risk who live alone considering the family size or household size is less than five members. Hence, our study revealed that household size (>5members) has lower odds of mortality risk compared to the household size (<=5 members).

Furthermore, our finding revealed that Central and Eastern region have significantly higher odds of communicable diseases related mortality while North-eastern has showed greater odds of diabetes-related mortality compared to Northern region. Though, higher hospitalization rate due to communicable diseases is depicted in the Central and Eastern regions among older adults in India (Akhtar & Saikia, 2022). A recent study has depicted that North-eastern region has higher premature mortality followed by Central and Eastern regions (Kumari & Mohanty, 2020).

### Strength and limitations of the study

The use of a newly available nationally representative sample gives robust cause-specific mortality estimates of the study variables and it is the first study which provides the association of cause-specific mortality with the socio-economic and demographic characteristics which is a major strength of this work. However, our study has several limitations. First, our study has used cross-sectional data from the LASI-(wave-I), therefore the causal comprehension is limited. Second, we have only included the top three disease-specific mortality variable in the study. Third, the self-rated health, educational attainment, income, and occupation status are well-known predictor of mortality across the life span but we have not included it in the study due to data unavailability. Furthermore, because there is a proportional relationship between physical-activity, dietary habits, lifestyle factors, obesity, tobacco use, smoking, and alcohol intake and ill health with mortality, we could not include these significant factors because of limited information of deceased adults were provided in the given data.

## Conclusions

Our study provides concrete evidence on the emerging socio-economic and demographic determinants among the 45+ adults which is significantly important to stress the urgent need for health planners and governments to intervene. To accomplish the Sustainable Development Goal (SDG), a quicker decline in premature NCD mortality death is needed to offset NCD fatalities caused by population aging, and global resources must be shifted to overcome the equity gap. The government should focus on preventing mass premature death and better preparing the world for future pandemics. Greater investments in health and promote healthy environments by addressing the common risk factors –lack of physical activity, tobacco, alcohol, air pollution and unhealthy diets – and ensuring that everyone who needs it has access to essential and lifesaving diagnosis, treatment, and care.

## Data Availability

The datasets generated and/or analysed during the current study are available with the International Institute for Population Sciences, Mumbai, India repository and could be accessed from the following link: https://iipsindia.ac.in/sites/default/files/LASI_DataRequestForm_0.pdf. Those who wish to download the data have to follow the above link. This link leads to a data request form designed by International Institute for Population Sciences. After completing the form, it should be mailed to: for further processing. After successfully sending the mail, individual will receive the data in a reasonable time.

## Acknowledgements

None

