## Supplemental File 1 for "Socio-economic and demographic determinants of all-cause, main-cause and sub-cause mortality among 45+ adults: Evidence from Longitudinal Ageing Study in India"

Supplementary file

CV026. What was the main cause of death?

1. Communicable, Maternal, Perinatal and Nutritional Conditions
   0. 1.1.  Tuberculosis (lungs, intestine, bones, brain)
   1. 1.2.  HIV/AIDS (Immune system)
   2. 1.3.  Diarrheal diseases (gastrointestinal)
   3. 1.4.  Malaria
   4. 1.5.  Other infectious and parasitic diseases
   5. 1.6.  Respiratory infections (Lungs and respiratory tract)
   6. 1.7.  Maternal conditions (related to pregnancy or post-delivery complications)
   7. 1.8.  Neonatal deaths
   8. 1.9.  Nutritional deficiencies (deficiency or excess of nutrients)
   9. 1.10.  Fever of unknown origin (despite investigations by a physician no explanation has been found)
2. Non-Communicable Diseases
   0. 2.1.  Malignant and other neoplasms (cancer)
   1. 2.2.  Diabetes mellitus
   2. 2.3.  Neuro-psychiatric conditions (brain)
   3. 2.4.  Cardiovascular diseases (heart)
   4. 2.5.  Chronic respiratory diseases (lungs)
   5. 2.6.  Diseases of the digestive system (gastrointestinal)
   6. 2.7.  Genitourinary diseases (genitals and urinary system)
   7. 2.8.  Musculoskeletal diseases (muscles and bones)
   8. 2.9.  Congenital anomalies (birth defects)
   9. 2.10.  Other Non-communicable diseases
3. Injuries
   0. 3.1.  Unintentional injuries: Motor vehicle accidents
   1. 3.2.  Unintentional injuries: Other than motor vehicle accidents
   2. 3.3.  Intentional injuries: Suicide
   3. 3.4.  Intentional injuries: Other than suicide
   4. 3.5.  Injuries of undetermined intent
4. Symptoms, Signs & Ill Defined Conditions
   0. 4.1.  Senility (related to old age)
   1. 4.2.  Ill- defined/All other symptoms, signs and abnormal clinical and laboratory findings
